# Evaluating the feasibility, sensitivity, and specificity of next-generation molecular methods for pleural infection diagnosis

**DOI:** 10.1101/2023.10.22.23297281

**Authors:** Peter T. Bell, Timothy Baird, John Goddard, Olusola S. Olagoke, Andrew Burke, Shradha Subedi, Tiana R. Davey, James Anderson, Derek S. Sarovich, Erin P. Price

**Author notes:** Authors contributed equally (Peter T. Bell, Timothy Baird, Derek S. Sarovich, Erin P. Price). Corresponding Author Erin P. Price University of Sunshine Coast.

## Abstract

**Rationale:** Pleural infections are common and associated with substantial healthcare cost, morbidity, and mortality. Accurate pleural infection diagnosis remains challenging due to low culture positivity rates, frequent polymicrobial involvement, and non-specific diagnostic biomarkers.

**Objective:** To undertake a prospective pilot study examining the feasibility and challenges associated with molecular methods for diagnosing suspected pleural infection.

**Methods:** We prospectively characterised 26 consecutive, clinically suspected pleural infections, and 10 consecutive control patients with suspected non-infective pleural effusions, using shotgun metagenomics, bacterial metataxonomics, quantitative PCR, and conventional culture.

**Results:** We demonstrate the feasibility of culture-independent molecular techniques for diagnosing suspected pleural infection. Molecular methods exhibited excellent diagnostic performance, with each method identifying 54% (14/26) positive cases among the pleural infection cohort, versus 38% (10/26) with culture. Meta-omics methods unveiled complex polymicrobial infections largely missed by culture. Dominant infecting microbes included streptococci (*S. intermedius*, *S. pyogenes*, *S. mitis*), *Prevotella* spp. (*P. oris*, *P. pleuritidis*), staphylococci (*S. aureus*, *S. saprophyticus*), and *Klebsiella pneumoniae.* However, we encountered challenges that complicated pleural infection interpretation, including: i) uncertainties regarding microbial pathogenicity and the impact of prior antibiotic therapy on diagnostic performance; ii) lack of a clinical diagnostic gold-standard for molecular performance comparisons; iii) potential accidental microbial contamination during specimen collection and processing; and iv) difficulties distinguishing background microbial noise from true microbial signal, particularly in low-biomass specimens.

**Conclusions:** Our pilot study demonstrates the potential utility and value of molecular methods in diagnosing pleural infection and highlights key concepts and challenges that should be addressed when designing larger prospective trials.

**Key messages:** *What is already known on this topic:* Confident pleural infection diagnosis is often challenging due to low culture positivity rates, frequent polymicrobial involvement, and non-specific diagnostic biomarkers. Limitations of conventional diagnostic tests result in prolonged and inappropriately broad-spectrum antimicrobial use, leading to potentially poorer patient outcomes and avoidable adverse effects.

*What this study adds:* We demonstrate the feasibility, utility, and challenges associated with the use of culture-independent molecular techniques for more accurate pleural infection diagnosis in a real-world clinical setting.

*How this study might affect research, practice, or policy:* These data will help to inform the design of larger prospective clinical trials and identify potential obstacles to be overcome as next-generation sequencing technologies become integrated into routine clinical practice.

## Introduction

Pleural infection is a complex, heterogeneous, and challenging clinical entity associated with prolonged hospitalization, significant healthcare costs, and high morbidity and mortality^1–4^. Clinical management frustrations arise due to insensitivities in conventional microbiologic diagnostics (culture-positive rates of only 20-60%^5–9^), resulting in prolonged and potentially inappropriate broad-spectrum antimicrobial use. Culture-based diagnostic sensitivity is poor due to multiple factors, including empiric antibiotic therapy prior to pleural sampling, low pathogen concentration in pleural fluid, and difficulties in cultivating fastidious microorganisms^4,10^. These limitations have obscured characterization of the pleural ‘microbiome’ (i.e. the entire microbial community), hampering clinical progress^11^.

Molecular diagnostics enable culture-independent microbial identification from clinical specimens^12^. Such techniques include shotgun metagenomics, metataxonomics, and quantitative PCR (qPCR), each having unique capabilities and drawbacks. Shotgun metagenomics involves total microbiome DNA sequencing of a sample, allowing strain-level detection of bacteria, archaea, fungi, parasites, and DNA viruses, along with their putative functions (e.g. antimicrobial resistance, virulence). However, metagenomics overlooks RNA viruses, and cannot readily differentiate living from dead microbes. Metataxonomics involves next-generation amplicon sequencing of conserved bacterial (16S ribosomal RNA [rRNA]) or fungal internal transcriber spacer (ITS) regions to characterize bacterial or fungal consortia, respectively; however, these methods typically only resolve microbiota to the genus level, and cannot be used for relative taxon quantification due to highly variable 16S rRNA/ITS copy numbers amongst species. Finally, qPCR is an inexpensive, same-day diagnostic method, with broad resolution ranging from strain to domain level; however, this method is typically limited to *a priori* target taxa^13,14^.

Several recent studies have applied culture-independent techniques to characterize pleural infections. This collective work has revealed a pleural microbiome that is often polymicrobial and characterized by a diverse range of classical pathogens and bystander microbes^7,8,15–21^. Earlier investigations implemented bacterial metataxonomics to profile the bacterial microbiome^7,8,15,21^; however, three recent studies have implemented metagenomics to more comprehensively profile pleural microbiota^16,19,20^. Xu *et al*. 2022 found bacterial dominance in 80 adult Chinese patients with pleural infections, most commonly *Streptococcus* and *Prevotella* spp., although fungi were occasionally identified^16^. However, the authors only reported taxonomic findings to the genus level, limiting in-depth understanding of microbiome composition and diversity^16^. Liu *et al*. 2023 reported on a single case study of viral pleurisy caused by Epstein-Barr virus, which was successfully treated with acyclovir^20^. Finally, Liu *et al*. 2023 examined microbiomes of pleural and ascitic fluid in 92 specimens retrieved from 32 Chinese children, identifying *Klebsiella pneumoniae* as the dominant pathogen (*n*=17), followed by *Escherichia coli* (*n*=9), and *Acinetobacter baumannii* (*n*=7); yeasts (*n*=6) and several viruses (*n*=33) were also identified^19^. In all three studies, metagenomics yielded a higher pathogen positive rate than conventional culture^16,19,20^.

To expand knowledge of the pleural microbiome in other cohorts, we undertook a prospective, observational, single-center pilot study of 26 consecutive patients presenting with suspected pleural infection at an Australian hospital, and 10 consecutive control patients lacking clinical suspicion of pleural infection. Next, we compared the diagnostic performance of microbe-enriched shotgun metagenomics, bacterial metataxonomics, panbacterial qPCR, and panfungal qPCR with conventional culture.

## Methods

### Ethics Statement

The study was approved by The Prince Charles Hospital Human Research Ethics Committee (HREC/2022/QPCH/81858) and the Sunshine Coast Hospital and Health Service Research Governance Office. All participants provided written informed consent.

### Study Participants

All patients ≥18 years of age presenting with suspected pleural infection (denoted with a ‘-P’ suffix) admitted between March and November 2022 to the Sunshine Coast University Hospital (SCUH), QLD, Australia, and undergoing a diagnostic and/or therapeutic procedure for suspected pleural infection, were prospectively invited.

Due to the lack of a ‘gold-standard’ pleural infection confirmation test, ‘clinical likelihood of pleural infection’ was objectively stratified by a specialist respiratory physician using predefined clinical criteria: 1. *Probable Pleural Infection*: (i) clinical evidence of pleural infection based on fever, elevated white cell count with left shift, pleurisy presence, raised inflammatory markers, and suitable clinical presentation; AND (ii) either direct evidence of pleural infection defined as the presence of Gram stain and/or culture positivity on pleural fluid; OR indirect evidence of pleural infection based upon pleural fluid pH <7.2 as measured on a blood gas analyser and/or glucose <3.0 mmol/L AND (iii) accompanied by at least two of the following criteria: serum C-reactive protein (CRP) >100 mg/L; complex pleural fluid and evidence of infection on imaging (pleural fluid loculation/septation on ultrasound and/or computed tomography); fever (>38.0°C). 2. *Possible Pleural Infection:* (i) clinical evidence of pleural infection (as defined in (i) above), AND (ii) negative Gram stain and culture negative on pleural fluid AND pH ≥7.2 and glucose >3.0 mmol/L) AND (iii) accompanied by at least 2 of the following criteria: CRP >100 mg/L; complex (i.e. loculated) pleural fluid collection on imaging; fever (≥38.0 °C). 3*. Unlikely pleural infection:* (i) No clinical evidence of pleural infection, AND (ii) all of the following criteria are met: pleural fluid Gram stain and culture negative with pH ≥7.2, glucose >3.0 mmol/L, serum CRP <100 mg/L, uncomplicated effusion on CT and or ultrasonographic imaging, and an alternate diagnosis confirmed (i.e. heart failure, malignancy). Based on these criteria, 26 consecutive patients with clinically suspected pleural infections were recruited.

For the control group, patients with pleural effusions without clinical suspicion for pleural infection at the time of sampling (denoted with a ‘-C’ suffix), were enrolled. These individuals were approached if they had an alternative effusion cause (e.g., malignancy or heart failure), absence of clinical history to support active infection, and simple effusion appearance on ultrasound and/or computed tomography. Patients were excluded if they had prior sampling of the pleural space, any clinical concern for current or recent infection, concomitant abdominal ascites, hepatic hydrothorax, or antibiotic administration within 3 weeks of pleural sampling. Ten participants meeting these criteria were enrolled as non-infected control subjects.

### Specimen Collection, Storage, and Processing

The study scientific and pathology teams were provided participant categorisation by the study clinicians (‘-P’ vs. ‘-C’) but no other information. For the pleural infection cohort, specimens were collected as soon as practicable using aseptic technique. Antibiotic commencement ranged from 21 days prior to pleural sampling to immediately post-pleural aspiration (**Table S1**). Following 10mL 1% lidocaine subcutaneous administration, between 10 and 60mL pleural fluid was collected into two sterile specimen pots. One pot was immediately mixed with 7mL stabilization agent^22^ for total DNA extraction (bacterial metataxonomics and qPCR); the second pot contained only pleural fluid for culture and host DNA depletion and extraction (metagenomics). Specimens were immediately stored at 4°C until processed. In parallel, specimens were also collected for routine microscopy (cell count, white blood cell differential, Gram stain), and biochemistry analyses (pleural fluid pH, protein, glucose, lactate dehydrogenase; serum CRP).

Microbial culturing was undertaken by both SCUH’s onsite Pathology laboratory and our Research laboratory, with VITEK 2 (bioMérieux, NSW, Australia) or Illumina NovaSeq 6000 whole-genome sequencing (WGS; Australian Centre for Ecogenomics, QLD, Australia) used respectively to confirm species identity. Bacterial metataxonomics was carried out using Illumina MiSeq 300bp V3-V4 16S rRNA amplicon paired-end sequencing^23^ (Ramaciotti Centre for Genomics, NSW, Australia). Metagenomic sequencing of host-depleted DNA^24^ was performed using Illumina NovaSeq 6000 150bp paired-end reads (17-27 million reads/sample; Azenta Life Sciences, China). For qPCR analyses, total genomic DNA was tested against panbacterial (16S rRNA)^25^, panfungal (28S rRNA)^26^, and human (β-globin)^27^ qPCR assays to quantify microbial-to-host load. For the panbacterial qPCR assay, a cycles-to-threshold (*C*_T_) cut-off value of 31 was used to delineate pleural infection-positive versus -negative cases. Further methodological details are described in Supplementary Methods.

### Statistical Analyses

To determine diagnostic test performance, ‘*Probable Pleural Infection’* was used as the infection-positive reference. Specificity, sensitivity, positive predictive value (PPV), and negative predictive value (NPV) were calculated for microbial culture, qPCR, bacterial metataxonomics, and metagenomics. Spearman’s rank correlation (*r_s_*) was used to determine pleural infection/no-infection diagnostic correlations between the four testing methods.

## Results

### Study participants and clinical outcomes

Twenty-six participants (*n=*7 female; median age=67 years [range 20 – 95]) with suspected pleural infection had an exudative pleural effusion as defined by Light’s criteria^28^. Twenty-four (92%) had loculated (complex) pleural effusions on ultrasound and/or computed tomography imaging, with 20 (77%) also having associated lung consolidation at the time of cross-sectional imaging. Twenty-five (96%) underwent intercostal catheter insertion, whereas one underwent diagnostic/therapeutic needle aspiration. All 26 patients received broad-spectrum antimicrobial therapy (**Table S1)**. Intrapleural enzyme therapy (5mg DNase and 10 mg tissue plasminogen activator) was administered in 17 participants (65%), and four (15%) required surgical decortication (**Table 1 & Table S2**). Thirty-day mortality was 12% (*n*=3).

**Table 1.** Clinical, Demographic and Outcome Data.

|  | **Suspected**<br>**Pleural Infection** | **Non-infectious Pleural**<br>**Effusion (Control)** |
| --- | --- | --- |
| ***Clinical / Demographics*** |  |  |
| Number of participants in group, *n* | 26 | 10 |
| Age in years (median, range) | 67 (20-95) | 78 (41-87) |
| Female sex, *n* (%) | 7 (27) | 2 (20) |
| Immunosuppression, *n* (%) | 3 (12) | 1 (10) |
| Fever, *n* (%) | 23 (90) | – |
| ICU admission, *n* (%) | 6 (23) | – |
| ***Biochemical*** |  |  |
| Serum White Cell count, cells x10<sup>9</sup>/L (median, range) | 14.5 (8.4-33.6) | 8.3 (4.3-15.2) |
| Serum C-reactive protein, mg/L (median, range) | 193 (39-547) | 56 (5-124) |
| Serum Procalcitonin, µg/L (median, range) | 0.6 (0.1-9.6) | 0.2 (0.1-0.3) |
| Serum LDH, unit/L (median, range) | 236 (108-477) | 208 (160-391) |
| Serum Protein, g/L (median, range) | 61 (49-76) | 62 (45-72) |
| Fluid LDH, unit/L (median, range) | 865 (173-17400) | 239 (81-672) |
| Fluid Protein, g/L (median, range) | 46 (23-58) | 43 (37-46) |
| Fluid pH (median, range) | 7.0 (6.6-7.5) | 7.4 (7.3-7.5) |
| Fluid glucose, mmol/L (median, range) | 3.9 (2.4-11.9) | 6.3 (4.6-10.0) |
| ***Imaging*** |  |  |
| Consolidation, *n* (%) | 20 (80) | – |
| Complex appearance of effusion, *n* (%) | 24 (92%) | – |
| ***Outcome Data*** |  |  |
| IPE, *n* (%) | 17 (65) | – |
| CTS, *n* (%) | 4 (15) | – |
| 30-day mortality, *n* (%) | 3 (12) | – |
| Pleural infection Likelihood* (%) | 1=0%, 2=38.5%,<br>3=61.5% | 1=100%, 2=0%, 3= 0% |

*Abbreviations:* Cardiothoracic Surgical decortication; IPE, Intrapleural Enzyme therapy; LDH, Lactate dehydrogenase. \* 1 = *Unlikely pleural infection*, 2 = *Possible pleural infection*: 3 = *Probable pleural* *infection*.

Among the 10 control subjects, 9 were diagnosed with malignant pleural effusion (MPE), and one with decompensated congestive heart failure (**Table 1 & Table S2**). Thirty-day mortality was not recorded for this cohort.

### Conventional culture performance

Pathology lab testing returned 10 (38%) microbial culture-positive results from the pleural infection cohort (**Table 2**), whereas only six (23%) were culture-positive from Research lab testing; however, there was 100% overlap with the Pathology results. Unexpectedly, pleural fluid from two control participants, SCHI0152-C and SCHI0175-C, cultured *Candida parapsilosis* and *Moraxella osloensis*, respectively, in the Research lab. No microbes were cultured in the control cohort by the Pathology lab.

**Table 2.**
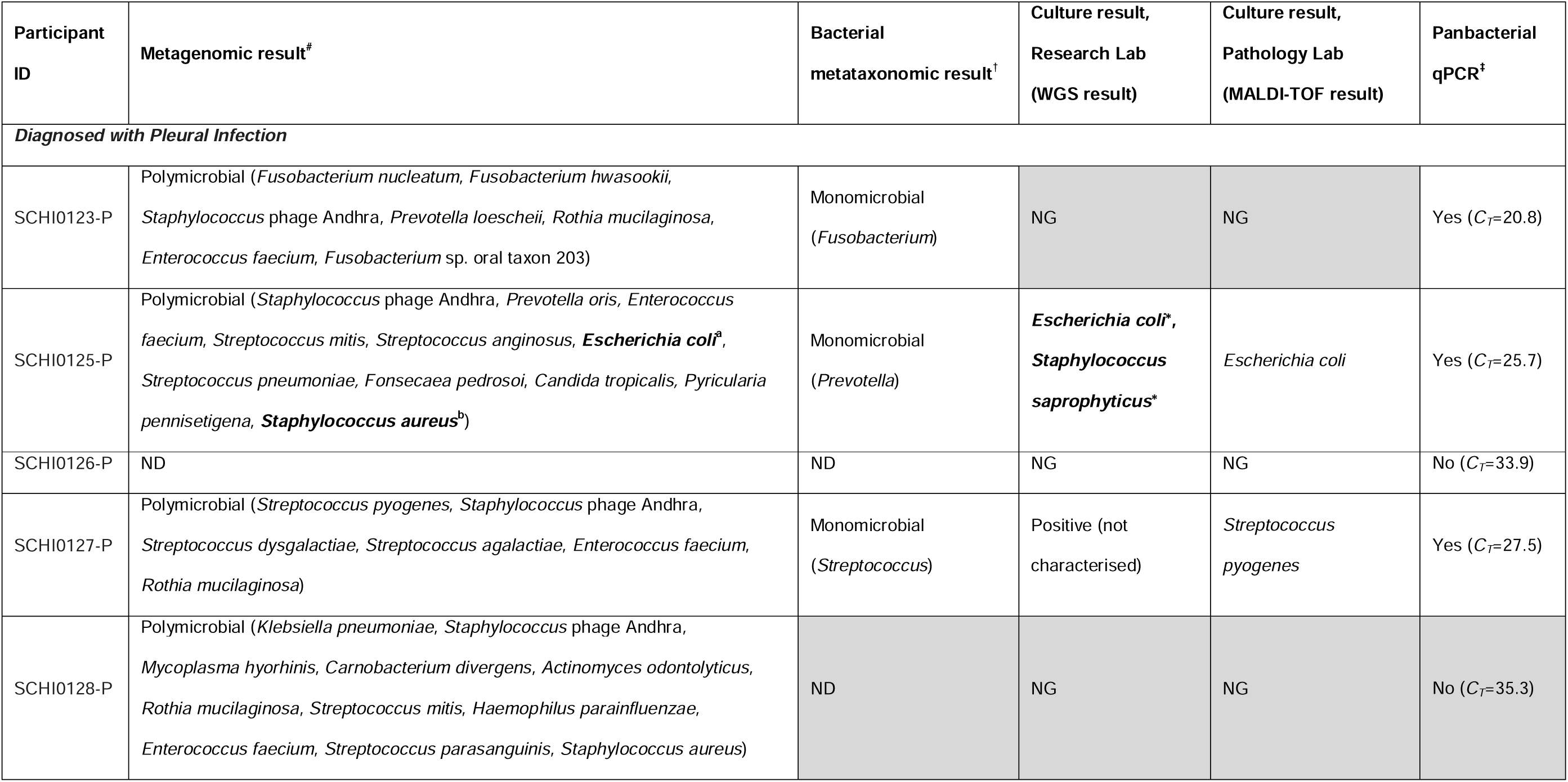
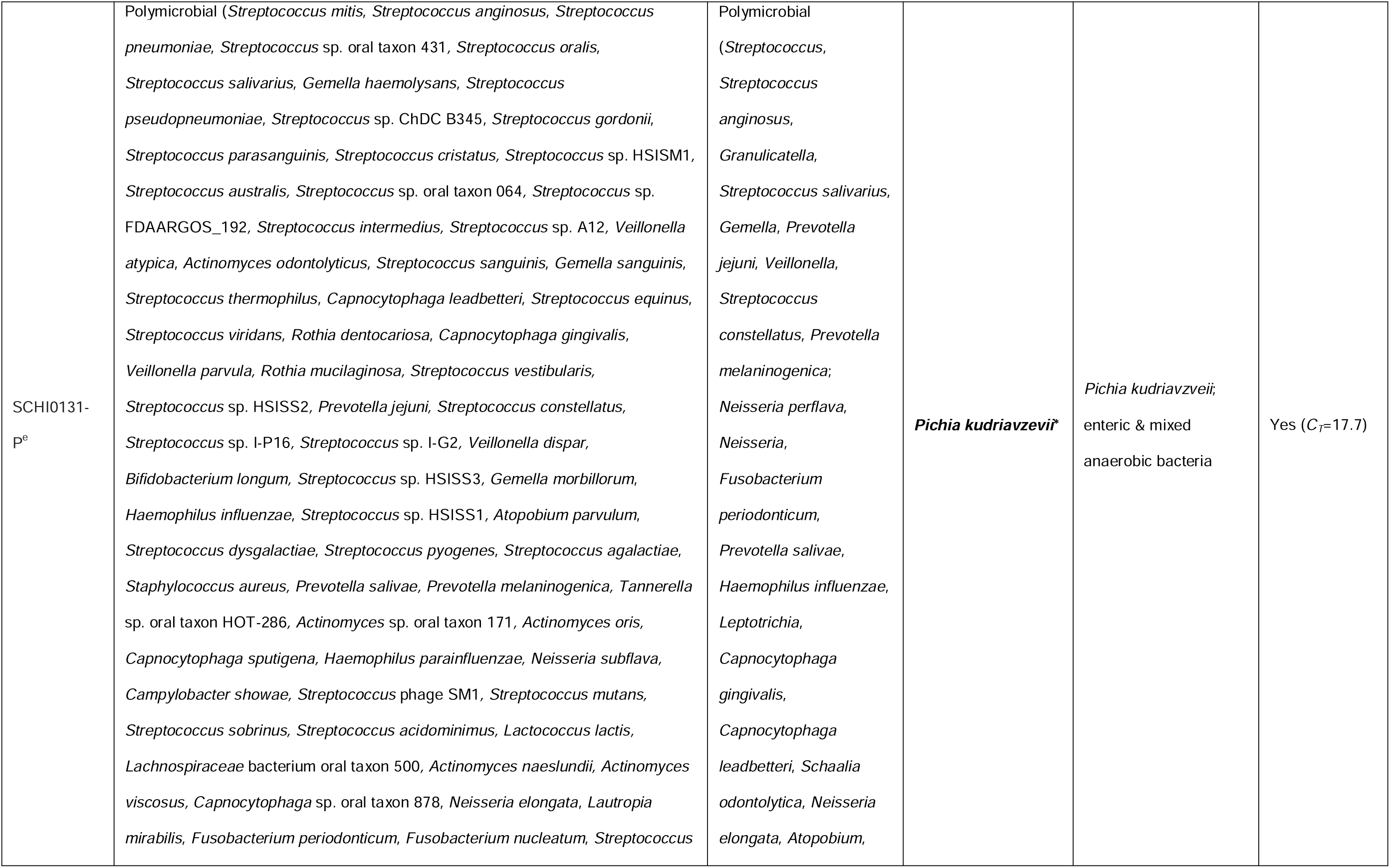
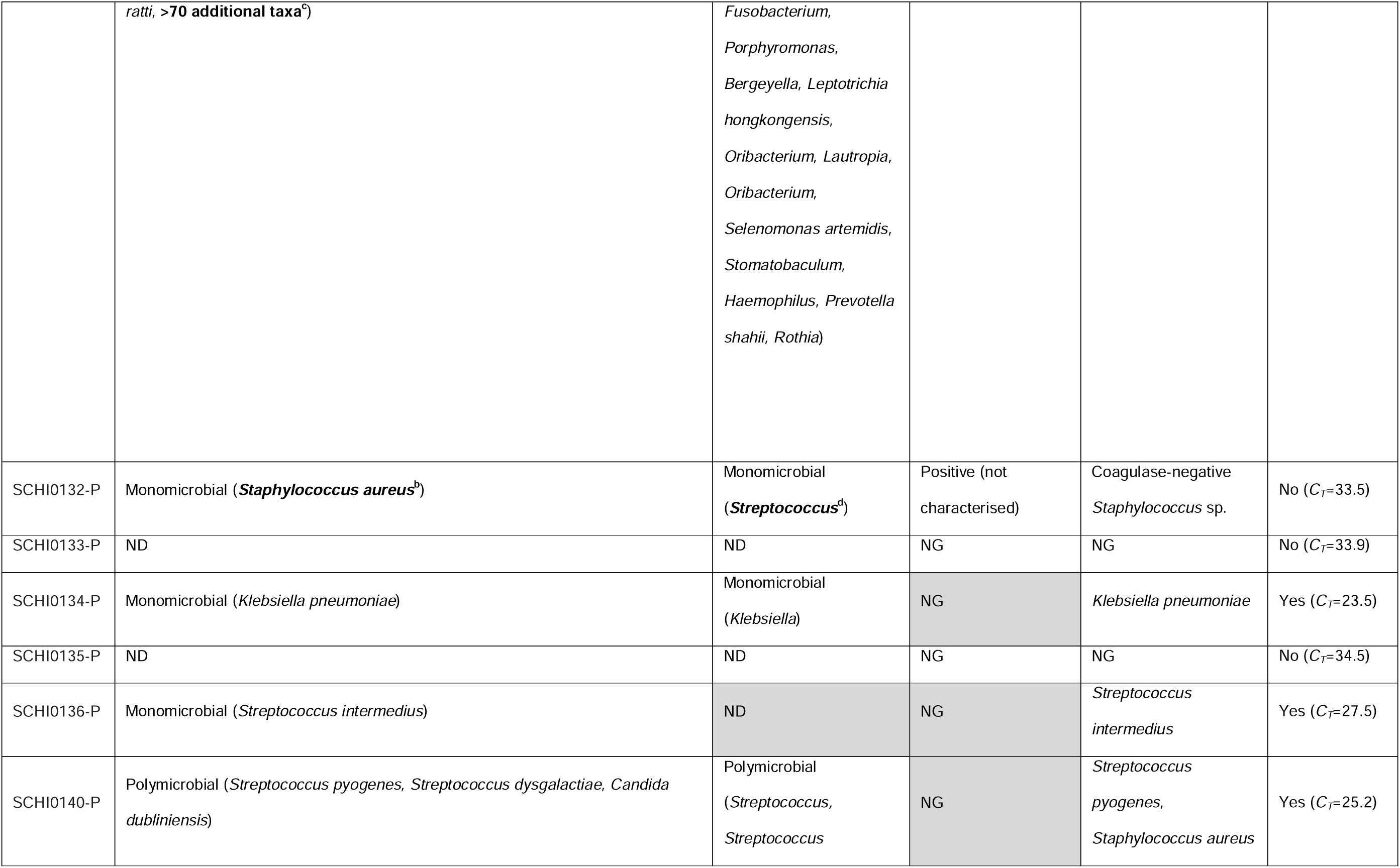
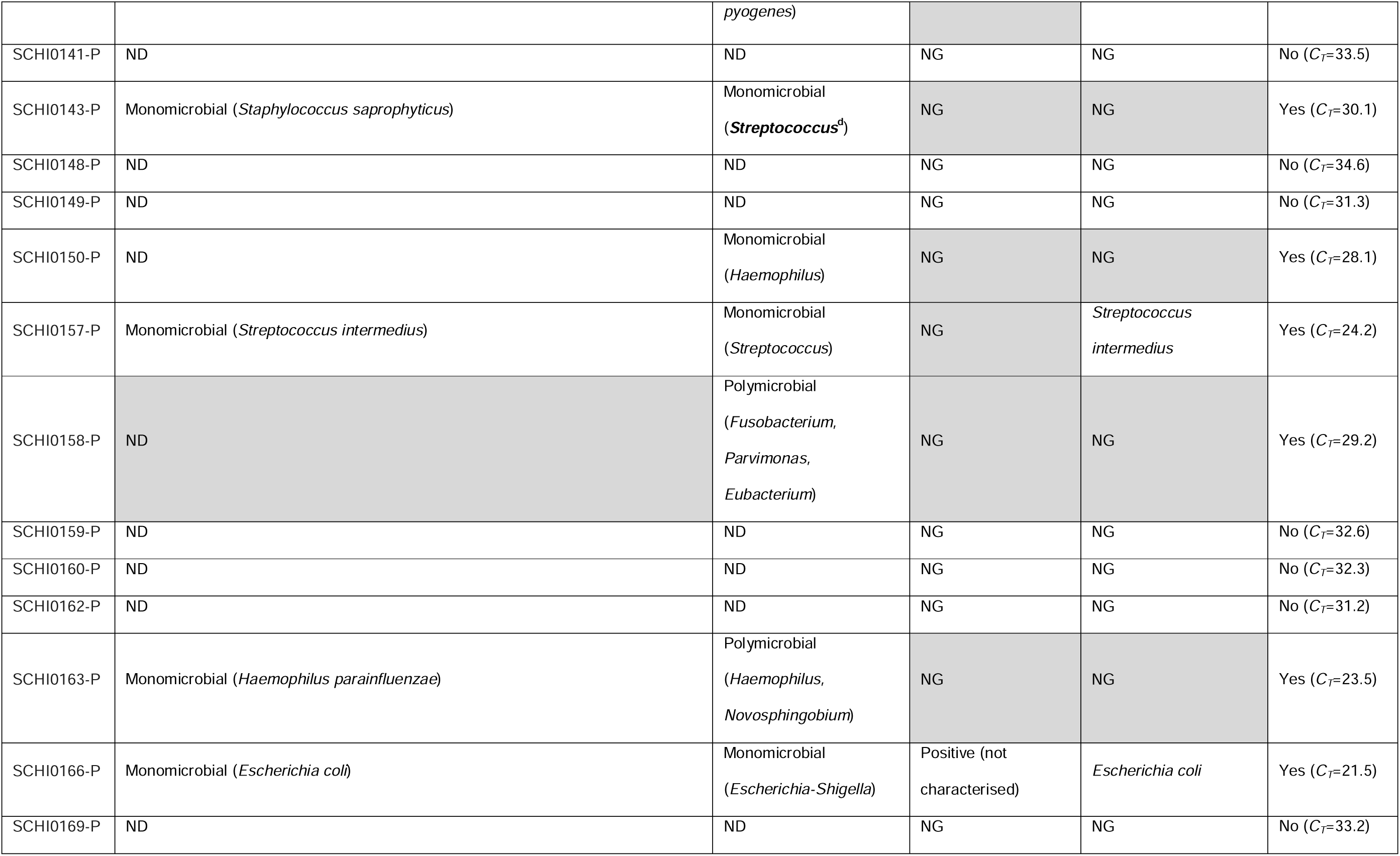
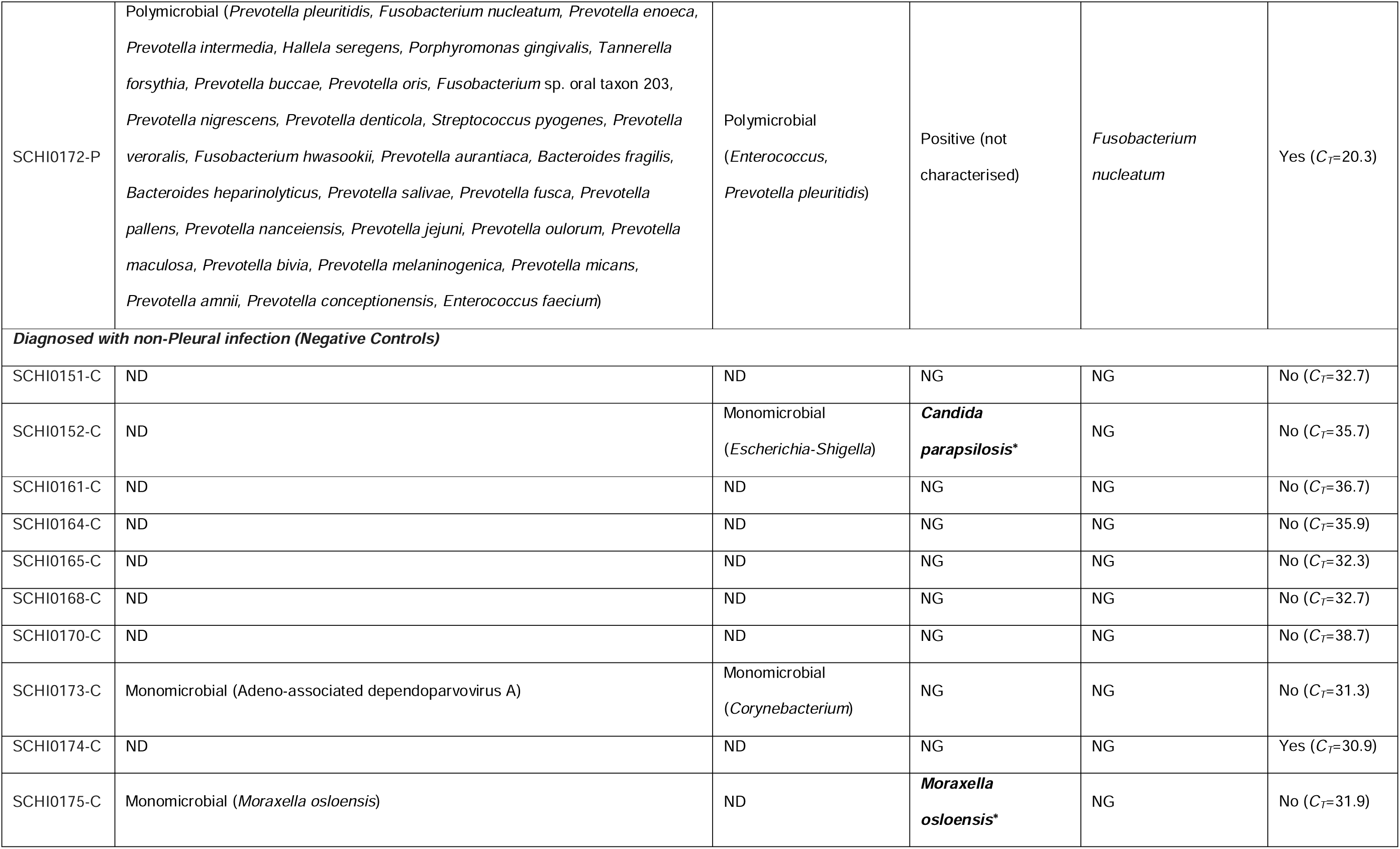
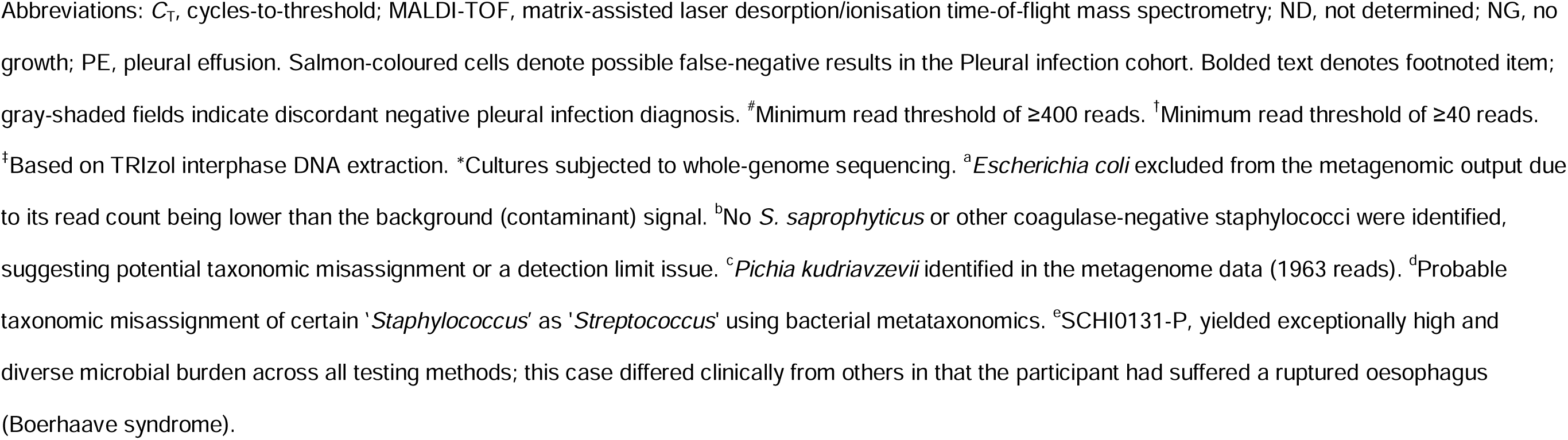
Pleural space diagnostic performance across 26 pleural infection diagnosed participants and 10 controls with a diagnosed non-pleural infection aetiology.

### qPCR performance

Positive bacterial detection was achieved with panbacterial qPCR in 14/26 (54%) suspect pleural infection cases (**Table 2**). All panbacterial qPCR-positive results were also bacteria-positive according to metagenomics and/or bacterial metataxonomics (**Table 2**). One of the 10 control cases was panbacterial qPCR-positive, albeit at a borderline *C_T_* value of 30.9; this result was not supported by any of the other tested methods. Panfungal qPCR did not identify fungi in any pleural infection or control cohort specimens.

### Bacterial metataxonomic and shotgun metagenomic performance

Together, bacterial metataxonomics and metagenomics identified 16/26 (62%) suspect pleural infection cases as infection-positive, with 14/26 (54%) cases determined by each individual method. Metagenomics identified 7/14 (50%) cases as polymicrobial, vs. 5/14 (36%) polymicrobial cases using bacterial metataxonomics (**Table 2 & Figure 1**). Despite both methods identifying 14 pleural infection cases each, only 12 overlapped, and only three of these cases were identified as polymicrobial by both methods (**Table 2**). Far fewer taxa were identified by bacterial metataxonomics in two of these three cases (SCHI0131-P, SCHI0172-P) (**Table 2**).

**Figure 1.**
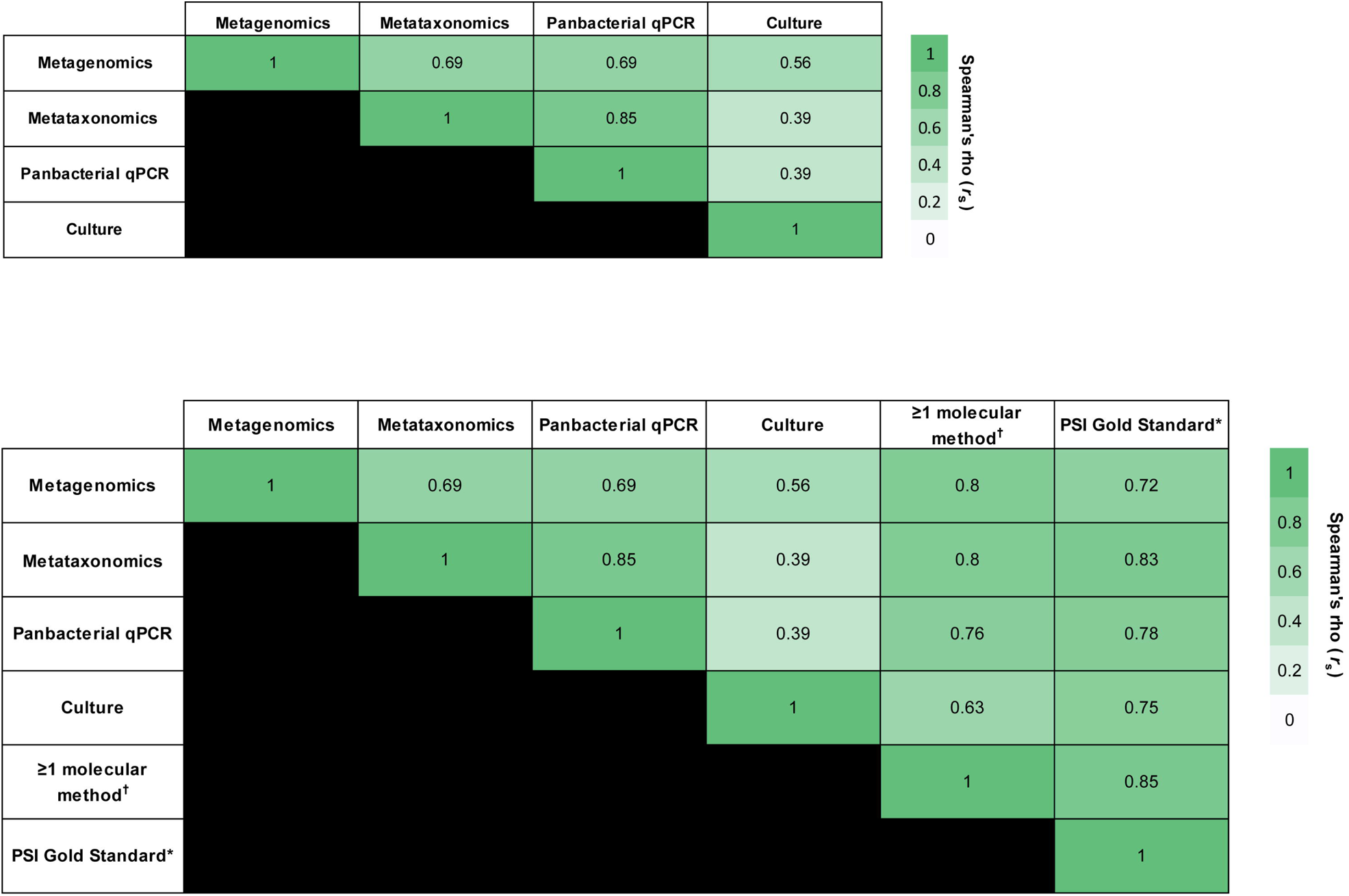
Metagenomic profiles identified from 26 suspected pleural infection specimens and 10 control cohort specimens. *Conventional culture positive; ^16S ribosomal RNA (panbacterial) quantitative PCR positive; #Whole-genome sequencing-confirmed (from culture). Dominant taxa are labelled on each bar.

### Molecular diagnostics versus microbial culture performance

Compared with our ‘pleural infection-positive’ clinical reference classification, all molecular diagnostic methods demonstrated superior sensitivity and NPVs compared with microbial culture (**Table 3**). As expected, specificity and PPVs remained high across all diagnostic methods.

**Table 3.** Comparative diagnostic accuracy of conventional culture and molecular methods for detecting pleural infection when compared to a surrogate gold-standard pleural infection classification of ‘probable pleural infection’*.

|  | **CONVENTIONAL**<br>**CULTURE** | **PANBACTERIAL**<br>**QPCR** | **BACTERIAL**<br>**METATAXONOMICS** | **METAGENOMICS** | **COMBINED**<br>**MOLECULAR** |
| --- | --- | --- | --- | --- | --- |
| **SENSITIVITY**<br>**(95% CI)** | 70.6 (44.0-89.7) | 82.4 (56.6-96.2) | 88.2 (63.6-98.5) | 82.4 (56.6-96.2) | 100 (80.5-100) |
| **SPECIFICITY**<br>**(95% CI)** | 100 (82.3-100) | 94.7 (74.0-99.9) | 94.7 (74.0-99.9) | 89.5 (66.9-98.7) | 84.2 (60.4-96.6) |
| **PPV**<br>**(95% CI)** | 100 (73.5-100) | 93.3 (68.1-99.8) | 93.8 (69.8-99.8) | 87.5 (61.7-98.4) | 85 (62.1-96.8) |
| **NPV**<br>**(95% CI)** | 79.2 (57.8-92.9) | 85.7 (63.7-97.0) | 90 (68.3-98.8) | 85 (62.1-96.8) | 100 (79.4-100) |

*Abbreviations:* CI, confidence interval; NPV, negative predictive value; PPV, positive predictive value; qPCR, quantitative PCR; Combined Molecular, all molecular methods (qPCR, bacterial metataxonomics, metagenomics). *\*Probable Pleural Infection:* Gram stain and/or culture positive on pleural fluid; or pleural fluid pH < 7.2 and 2 or more of the following: serum C-reactive protein > 100 mg/L; complex pleural fluid on imaging; fever (> 38°C)

Conventional culture somewhat correlated with molecular methods (metagenomics: *r_s_*=0.56 [*p*<0.001]; bacterial metataxonomics: *r_s_*=0.39 [*p*=0.05]; panbacterial qPCR: *r_s_*=0.39 [*p*=0.05]) (**Figure 3**). Metagenomics demonstrated the strongest correlation across methods (bacterial metataxonomics: *r_s_*=0.69 [*p*<0.001]; panbacterial qPCR: *r_s_*=0.69 [*p*<0.001]; conventional culture: *r_s_*=0.56 [*p*<0.01]). The strongest individual correlation was between panbacterial qPCR and bacterial metataxonomics (*r_s_*=0.85; *p*<0.001; **Figure 3**); this result was expected as both methods target the 16S rRNA gene.

### Pleural infection aetiology

A diverse array of oro-naso-hypopharyngeal-, oral-, dental-, respiratory tract-, and gut-associated microbes were detected in the 14 metagenomic infection-positive specimens (**Table 2 & Figure 1**). Streptococci (*n*=5; 2 x *S. intermedius*, 2 x *S. pyogenes*, 1 x *S. mitis*) dominated, followed by *Prevotella* spp. (*n*=2; *P. oris* and *P. pleuritidis*), *Staphylococcus* spp. (*n*=2; *S. aureus* and *S. saprophyticus*), and *Klebsiella pneumoniae* (*n*=2) (**Table 2**). Seven (50%) pleural infection cases were monomicrobial according to metagenomics, of which two harboured *S. intermedius*, along with one each of *K. pneumoniae*, *S. saprophyticus*, *S. aureus*, *Enterococcus faecium*, and *Escherichia coli* (**Table 2 & Figure 2)**. In contrast, bacterial metataxonomics identified nine (35%) monomicrobial pleural infection cases, of which four were *Streptococcus* sp., along with one each of *Escherichia-Shigella* sp., *Fusobacterium* sp., *Haemophilus* sp., *Klebsiella* sp., and *Prevotella* sp. (**Table 2 & Figure 2**). Five monomicrobial cases overlapped with the two meta-omics methods; however, bacterial metataxonomics erroneously assigned two monomicrobial pleural infection cases, SCHI0132-P and SCHI0143-P, as ‘*Streptococcus*’, despite the SCHI0132-P isolate being confirmed as coagulase-negative *Staphylococcus* sp. by VITEK 2 (**Table 2**), indicating a probable bacterial metataxonomic database error for this genus.

**Figure 2.**
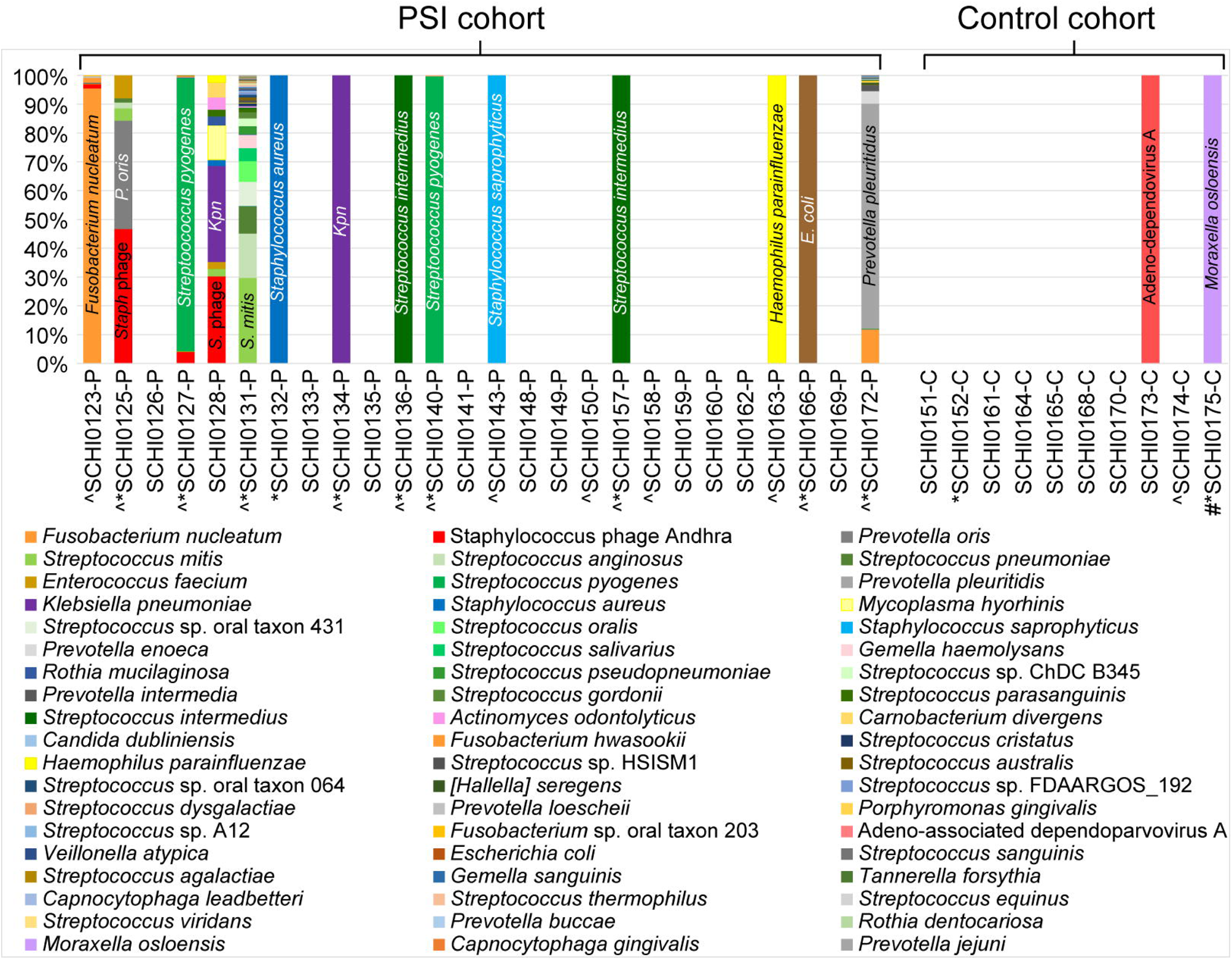
Bacterial 16S ribosomal RNA (rRNA) metataxonomic profiles identified from 26 suspected pleural infection specimens and 10 control cohort specimens. *Conventional culture positive; ^16S ribosomal RNA (panbacterial) quantitative PCR positive; #Whole-genome sequencing result from culture; ‡Likely incorrect assignment of *Staphylococcus* as *Streptococcus* (1 case confirmed as a coagulase-negative *Staphylococcus* by MALDI-TOF analysis of culture). Dominant taxa are labelled on each bar.

**Figure 3.**
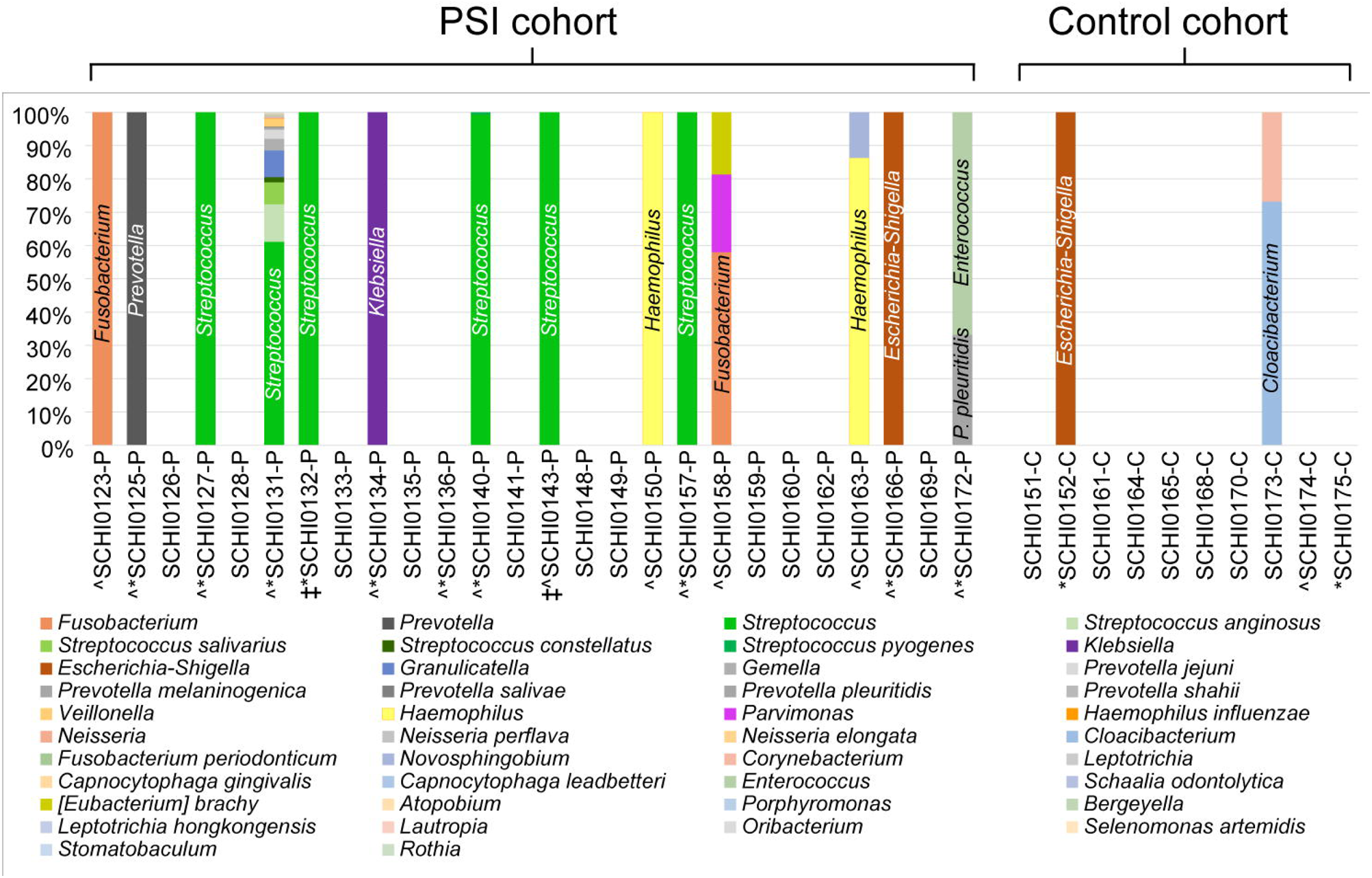
Correlation matrix of the pleural infection diagnostic methods used in this study. 1=perfect correlation; -1=no correlation. Conventional culture showed the weakest correlation when compared with each of the three molecular methods.

As expected, bacterial metataxonomics failed to identify fungi or viruses in the pleural infection cases, whereas conventional culture and metagenomics both identified the yeast, *Pichia kudriavzevii*, in SCHI0131-P. Metagenomics further identified the fungi *Rhizoctonia solani*, *Wallemia mellicola*, *Meyerozyma guilliermondii*, *Truncatella angustata*, *Cokeromyces recurvatus*, *Bipolaris maydis*, *Candida orthopsilosis*, and *Gilbertella persicaria* in SCHI0131-P (subject suffered from confirmed Boorhaaves syndrome), *Candida dubliniensis* in SCHI0140-P, and *Candida tropicalis*, *Fonsecaea pedrosoi*, and *Pyricularia pennisetigena* in SCHI0125-P. In all instances, fungi were in relatively low abundance. Metagenomics also identified bacteriophages in 5/26 infection-positive cases, with *Staphylococcus* phage Andhra seen in SCHI0123-P, SCHI0125-P, SCHI0127-P, and SCHI0128-P, and *Streptococcus* phage SM1 seen in SCHI0131-P. *Staphylococcus* phage Andhra was among the most dominant taxa seen in SCHI0125-P and SCHI0128-P, indicating very high phage levels in pleural fluid; both specimens harboured *S. aureus* according to metagenomics, and SCHI0125-P also had *S. saprophyticus* according to culture and WGS (**Table 2 & Figure 2**).

Metagenomics unexpectedly found microbes in 2/10 control cases (**Table 2**). The non-pathogenic DNA virus, adeno-associated dependoparvovirus A, was identified in SCHI0173-C – although no helper virus was observed – and *M. osloensis* was confirmed in SCHI0175-C. *C. parapsilosis* cultured from SCHI0152-C was not detected, suggesting possible operator contamination during specimen collection or processing. Bacterial metataxonomics failed to identify the culture-confirmed *M. osloensis* in SCHI0175-C, but identified *Escherichia-Shigella* in SCHI0152-C (1453 reads) and *Corynebacterium* in SCHI0173-C (190 reads), findings that were not supported by the other methods, again likely due to contamination.

### Microbiology and clinical outcomes

Of the three pleural infection cases with a <30-day mortality, two (SCHI0128-P and SCHI0134-P) were infected with *K. pneumoniae*, and two (SCHI0131-P and SCHI0128-P) had polymicrobial infections according to metagenomics. In contrast, no patient surviving beyond 30 days (*n*=23) harbored *Klebsiella* spp. according to any testing method.

### Antimicrobial resistance gene detection

*In silico* antimicrobial resistance gene detection only identified innate drug resistance genes, with no evidence of acquired resistance development in any case (**Table S1**).

## Discussion

Clinical decision-making in suspected pleural infection is challenging due to low culture positivity rates^5–8^, resulting in frequent diagnostic uncertainty and antimicrobial overuse. Recent work^7,8,15,16,19^ has highlighted potential advantages of molecular methods for diagnosing and determining the aetiology of pleural infection. Our pilot feasibility study adds to this growing body of work, being one of the first studies to apply shotgun metagenomics on pleural infections, and the first to prospectively collect and profile consecutive pleural fluid specimens. Our findings suggest that all three culture-independent molecular techniques tested in our study – metagenomics, bacterial metataxonomics, and panbacterial qPCR – have excellent performance characteristics for pleural infection diagnosis, and may be superior to conventional culture for diagnosis and aetiologic confirmation. Panbacterial qPCR stands out as a cost-effective, simple, and same-day diagnostic that could be readily implemented as a routine test for pleural fluid specimens; such a test may assist with both antimicrobial stewardship efforts and rapid treatment decision-making^29^. In support of its clinical utility, panbacterial qPCR showed excellent diagnostic performance (sensitivity 82%, specificity 95%, PPV 93%, NPV 86%) and strong correlation with shotgun metagenomics, bacterial metataxonomics, and conventional culture.

In line with recent molecular studies^7,8,15–17^, we confirm that pleural infections can be either monomicrobial or polymicrobial, with predominant microbes most likely originating from oral, dental, or upper and lower respiratory tract sources, and occasional incursion from gut-borne pathogens (e.g. *E. coli*, *Enterococcus faecium*, *K. pneumoniae*) (Table 2). Furthermore, metagenomics and bacterial metataxonomics unveiled greater microbial heterogeneity than could be detected by conventional culture. Similar to recent work^16,19^, we observed that polymicrobial bacterial infection was also occasionally associated with fungal coinfection (albeit in very low abundance) using metagenomics, confirming the challenging nature of diagnosing and treating complex polymicrobial pleural infections across microbial domains. Streptococcal and staphylococcal phages were also seen in five cases of pleural infection, with their host species not always detected, suggesting possible lytic activity towards their host within the pleural space, or incidental phage transit from other body sites^30^. Although not examined here, others have shown that phage presence in clinical specimens can negatively impact culture rates^31^. Despite low sample numbers, we found that *K. pneumoniae* presence was associated with greater risk of mortality, consistent with recent bacterial metataxonomic study that showed an association between Enterobacteriaceae and mortality^8^.

Immediate antimicrobial administration upon suspicion of pleural infection is important for preventing sepsis^32^; however, this practice may negatively affect pleural infection diagnostic rates. Despite this, many prior pleural microbiome studies have identified 100% infection positivity using molecular testing^7,8,15^. In contrast, only 62% of suspected pleural infections in our study had detectable microbes according to molecular and/or culture assessment. Our lower positivity rate likely reflects our prospective, consecutive recruitment of participants without prior knowledge of culture results or pleural biochemistry, an approach that captures the diagnostic uncertainty facing clinicians in real-world practice. Only two other studies have not used prior knowledge of pleural fluid biochemistry or microbiology for molecular pleural infection identification and characterisation; their infection positivity rate was 70%^16,17^. Our finding that 38% of suspected pleural infection patients did not have detectable microbes according to any method suggests that a subset of patients with clinically suspected pleural infection may instead have sterile pleural effusions at the time of sampling. In support of this conclusion, these patients had more benign pleural biochemistry using recognised cut-off values (i.e. pH <7.2; glucose <3.0 mmol/L)^1^ (**Table 1**). A second possibility is that some infections were missed due to prior antibiotic exposure; however, others have shown that molecular approaches are less impacted by prior antibiotics than conventional culture^7,16^. A third possibility is that human DNA depletion prior to metagenomic sequencing may have limited our ability to detect extracellular microbial DNA^24^. To overcome this issue, deeper metagenomic sequencing on total DNA can be conducted; however, such an approach is costly, and risks missing low-abundance microbes due to overwhelming human DNA signal^24,33^. Prospective randomized trials are needed to understand whether a truncated antibiotic course can be safely used in people with likely sterile effusions as identified with molecular diagnostics, and to quantify the impact of prior antibiotic administration on culture or molecular detection sensitivity.

Sensitive molecular assays come with the trade-off that microorganism presence might represent either a contaminant or a bona fide pathogen. To assess this issue, we investigated whether the control group, predominantly comprised of MPEs, had a pleural microbiome. Although most controls had a sterile pleural space, one MPE patient was positive for *Moraxella osloensis* on culture, WGS, and metagenomics. *M. osloensis*, a commensal of human skin and the upper respiratory tract, most commonly causes infection in cancer patients^34^; it is thus feasible that this patient had an undiagnosed infection alongside their MPE. Three other MPE patients also unexpectedly yielded microbes: *C. parapsilosis* was cultured from SCHI0152-C, and bacterial metataxonomics identified *Corynebacterium* and *Escherichia-Shigella* in SCHI0173-C and SCHI0152-C, respectively; however, these findings were not supported by the other methods. Although specimen or sampling contamination represents the most plausible explanation for these results, a recent bacterial metataxonomic study suggested that non-infective pleural disease may contain unique microbial signatures^7,35^. Strict aseptic practice during specimen collection and processing can mitigate some of these false-positive cases. However, microbial contamination may still occur at many stages of sample handling and analysis, including procedural contamination (e.g. remnant microbial DNA in the specimen jars or local anaesthetics), or residual DNA from transient microbial seeding of the pleura. In addition, index hopping and ‘kitome’ contamination are known drawbacks of next-generation sequencing, especially in low-biomass specimens like pleural fluid^33^. Careful consideration of these challenges will be important in the design of larger prospective studies. Further studies examining the pleural microbiome across a spectrum of pleural diseases, including in people with MPE and congestive heart failure, will also be illuminating.

Other challenges also complicated interpretation of pleural diagnosis and aetiology, including difficulties distinguishing background noise from true microbial signal, lack of appropriate methodology for detecting RNA viruses, and uncertainty regarding microbial causality and pathogenicity (e.g. the role of bystander microbes in pleural disease development and prognosis), particularly in polymicrobial infections. Shotgun metatranscriptomics, which involves total RNA sequencing of microbiome transcripts, may address these difficulties. However, to our knowledge, this technique has not yet been undertaken to investigate pleural infections, so its value in this cohort remains unclear. Perhaps most challenging of all is the current lack of gold-standard clinical diagnostic criteria for pleural infection. In clinical practice, an integrated approach is taken to weigh multiple factors to make a probabilistic diagnosis, including pleural biochemistry, host response, and imaging. In this study, we attempted to standardize integrated probabilistic diagnostic classification by formulating a pragmatic reference standard to compare diagnostic performance between modalities. However, the issue of a validated diagnostic gold-standard remains an unresolved challenge. It is also important to consider the impact of incorporation bias^36^ when interpreting the performance of conventional culture results, given that such results contribute to the diagnostic ‘reference standard’, thereby inflating culture performance estimates.

In conclusion, this prospective study demonstrates the feasibility of, and challenges associated with, molecular tests for pleural infection diagnosis. Whilst such methods hold great clinical utility and potential superiority in routine pleural infection diagnostics, questions and challenges remain. A large, rigorous study comparing molecular and culture-based diagnostic techniques in pleural infection is now required to further address these opportunities for diagnostic advancement, along with the associated challenges in eventual clinical translation.

## Author Contributions

The authors confirm contribution to the paper as follows: study conception and design, PTB, TB, JG, OSS, AB, DSS, EPP; data collection, PTB, TB, JG, OSS, AB, JA, DSS, EPP; analysis and interpretation of results, PTB, TB JG, OSS, AB, SS, TRD, DSS, EPP; draft manuscript, PTB, TB, DSS, EPP; preparation of manuscript, PTB, JG, OSS, AB, SS, TRD, DSS, EPP. All authors reviewed the results and approved the final version of the manuscript.

## Data Availability

All sequence data generated in this study are available via NCBI BioProject PRJNA972883; genome assemblies are available via NCBI BioProject PRJNA970939.

https://www.ncbi.nlm.nih.gov/bioproject/?term=PRJNA972883

https://www.ncbi.nlm.nih.gov/bioproject/?term=PRJNA970939

## Acknowledgements

We thank Emma Seaniger and Louise McIntosh for assistance with sample collection and study coordination, Michael Caffery and the Pathology Queensland laboratory scientists for their assistance with culture, and the Australian BioCommons team (led by Ashley Dungan at University of Melbourne, and Melissa Burke at University of Queensland) for providing QIIME 2 training.

