## Supplementary Methods for "Evaluating the feasibility, sensitivity, and specificity of next-generation molecular methods for pleural infection diagnosis"

***Pathology laboratory processing****.* Pathology laboratory testing involved two steps: i) inoculating ~10mL pleural fluid at the point of collection into anaerobic and aerobic blood culture bottles, which were incubated at 37^o^C for 5 days in a blood culture incubator (BacT/ALERT 3D, bioMérieux, North Ryde, NSW, Australia), and ii) direct plating of pleural fluid onto horse blood and chocolate agars, followed by 37^o^C incubation in 5% CO_2_ or anaerobic atmospheres for 5 days, with daily inspection for culture growth. All culture-positive specimens retrieved by the Pathology laboratory were identified using the VITEK MS (bioMérieux) matrix-assisted laser desorption/ionisation time-of-flight mass spectrometry system.

***Research laboratory processing****.* Pleural fluid was subjected to an initial centrifugation step of 290 x *g* at 4^o^C for 15 min to pellet gross cellular material (Pellet 1), as previously described^1^. The supernatant was transferred to a fresh tube and subjected to a second centrifugation step of 9200 x *g* at 4^o^C for 15 min to pellet remaining cellular material (Pellet 2). Approximately 20 µL of the resuspended pellets (for all specimens) and supernatant (for the first 10 PSI specimens) were subjected to heat soak DNA extraction using 5% chelex 100 ^2^ resin (Bio-Rad, South Granville, NSW, Australia) for 10-20 min at 95^o^C. Following resin pelleting at 10,000 *x* g for 1 min, supernatant was diluted 1:10 with water and used for quantitative PCR (see qPCR testing below). Following chelex qPCR testing, 2 x ~50μL samples of the best-performing fraction/s (Pellet 1, Pellet 2, or supernatant) were glycerol-stocked, and ~20μL specimen was plated and 16-streaked onto various agars (MacConkey, Sabouraud dextrose, mannitol salt, Columbia horse blood, and supplemented chocolate agar plates [Edwards Group, Narellan, NSW, Australia]) and incubated aerobically at 37^o^C to isolate common pathogens and oral/airway microbes. Blood and chocolate agars were also inoculated for anaerobic incubation in an Anaerobox jar (Thermo Fisher Scientific, Scoresby, VIC, Australia) using AnaeroPouch anaerobic gas generators (Thermo Fisher Scientific), with anaerobic atmosphere confirmed via resazurin anaerobic indicator strips (Thermo Fisher Scientific). Culture growth was assessed at 24h and then periodically for 7 days. Plates lacking any observable growth after 7 days were deemed as ‘no growth’.

***DNA extraction for metagenomic sequencing and whole-genome sequencing (WGS)****.* Pleural fluid was human DNA-depleted using the “Benzonase 1” method ^3^. Cells were then lysed using an in-house metapolyzyme concoction containing 50 U mutanolysin, 280 KU lysozyme, 4.4 U lysostaphin, 0.03 U chitinase, 10 U achromopeptidase (Sigma-Aldrich, Bayswater, VIC, Australia), and 60 U zymolyase (MP Biomedicals, Irvine, CA, USA), which was incubated in 200 µL enzymatic lysis buffer (1.2% Triton-X, 2X Tris-EDTA, pH=8.0) at 37^o^C for 2 hours. DNA extraction was carried out using the Gram-Positive Bacteria protocol from the DNeasy Blood & Tissue kit (Qiagen, Clayton, VIC, Australia). Seven reagent-only controls were included to account for background DNA contamination in extraction and sequencing reagents. Bacterial and fungal cultures for WGS were extracted as above, but without benzonase depletion.

***DNA extraction for bacterial metataxonomics***. The RNA stabilisation reagent specimens (excluding SCHI0123-P and SCHI0125-P) were centrifuged at 9200 x *g* and 4^o^C for 15 min to pellet all cellular material; the supernatant was decanted, followed by 10 parts addition of TRI Reagent and 30 µL β-mercaptoethanol (Sigma-Aldrich) to the pellet to inactivate RNases. For SCHI0123-P and SCHI0125-P, Pellet 2 from the pleural fluid pot was used for TRI Reagent DNA extraction due to no RNA pot being collected for these specimens. DNA was retrieved from the interphase/proteinaceous phase using the back extraction buffer method ^4^; the aqueous phase was removed and stored at -80^o^C for future RNA extraction. Three reagent-only controls were also processed.

***Bacterial metataxonomics****.* All 36 pleural fluid specimens were subjected to 16S rRNA V3-V4 amplification using Bakt_341f and Bakt_805r primers ^5^, followed by Illumina MiSeq 300bp paired-end sequencing (Ramaciotti Centre for Genomics, Sydney, NSW, Australia). Illumina adapters were demultiplexed and trimmed with QIIME 2 ^6^ using a *p* error rate of 0.20. DADA2 ^7^ was then used to denoise, quality-filter (forward and reverse reads trimmed to 261bp and 199bp, respectively), and assign amplicon sequence variants (ASVs) for taxonomic assignment using the silva-138-99-nb-classifier ^8^. A minimum cut-off of 40 reads was used to define microbial presence. Where at least one of the three reagent-only controls had a given taxon exceeding 40 reads, all PSI and control datasets had this equivalent signal manually removed from the tabulated barchart output. Only taxon assignments at the Genus or Genus-Species level were recorded; Phyla through Family levels were discarded. Raw bacterial 16S rRNA metataxonomic sequencing reads are available at NCBI BioProject PRJNA972883.

***Metagenomic Sequencing****.* Illumina 150bp paired-end reads (16-28 million reads/sample) were generated for all 36 specimens using the NovaSeq 6000 platform (Azenta Life Sciences, Suzhou, China), resulting in between 17-27 million reads per sample. Seven reagent-only controls were also sequenced to account for background microbial signal and index hopping. Metagenomic reads were first quality-filtered with Trimmomatic v0.39 ^9^ using TruSeq Illumina adapter removal, leading=3, trailing=3, sliding window=4 :15, and minimum length=36 ^10^. Next, human reads were identified using the Kraken 2 v2.1.2 ^11^ human database followed by removal using Seqtk v1.3 (<https://github.com/lh3/seqtk>). Non-human reads were assigned microbial taxonomies by interrogation against the default full Kraken 2 bacterial, fungal and viral database, with the fungal database expanded to include genomes of chromosome, scaffold and contig completeness. Contaminant signal, identified both from no-template controls and by manually assessing the ecology of each microbe, was removed using Seqtk. Microbes with higher abundance in one or more PSI specimens compared with no-template controls were retained. A minimum cut-off of 400 reads was used to define microbial presence in the clinical specimen metagenomic data. Human-depleted metagenomic reads are available at BioProject PRJNA972883.

***WGS****.* To confirm species identity, Illumina 100bp paired-end reads (34-496x) were generated for five cultures (SCHI0125.M.1, SCHI0125.M.2, SCHI0131.M.3, SCHI0152.S.1, and SCHI0175.S.1) retrieved from two PSI participants (SCHI0125-P and SCHI0131-P) and two control participants (SCHI0152-C and SCHI0175-C) using the NovaSeq 6000 platform (Australian Centre for Ecogenomics, St Lucia, QLD, Australia). Genomes were quality-filtered with Trimmomatic using the same parameters as the metagenome sequences, followed by *de novo* assembly with SPAdes v3.15.5 ^12^. Assemblies are available via BioProject PRJNA970939.

***AMR gene assessment*.** Human-depleted metagenomic reads were examined for AMR gene presence using ResFinder v4.1.11 ^13^. AMR genes present in no-template controls or metagenomic-negative samples, and exhibiting <100% identity, were excluded.
