## Supplementary material for "Evaluating the feasibility, sensitivity, and specificity of next-generation molecular methods for pleural infection diagnosis": Table S2

**Table S2. Demographic and clinical data of all participants examined in this study.**

| Clinical & Demographic Data | | | | | | Blood & Serum | | | | | Pleural Fluid | | | | Imaging | | Procedure | | Outcome | Pleural Infection  Likelihood* |
| --- | --- | --- | --- | --- | --- | --- | --- | --- | --- | --- | --- | --- | --- | --- | --- | --- | --- | --- | --- | --- |
| Participant ID^†^ | Age (5-year range) | Gender | immunosuppression | Fever | Intensive care unit admission | Total white cell  count (cells x10^9^/L) | C-reactive protein (mg/L) | Procalcitonin  (µg/L) | Serum LDH  (unit/L) | Serum Protein  (g/L) | Fluid LDH  (unit/L) | Fluid Protein  (g/L) | Fluid pH | Fluid Glucose  (mmol/L) | Consolidation | Nature of effusion | IPE | CTS | 30-day mortality | 1 – Unlikely  2 – Possible  3 –Probable/Definite |
| SCHI0123-P | 75-80 | M | 0 | 1 | 0 | 31 | 414 | 1.27 | 251 | 61 | 3310 | 53 | 6.6 | <0.3 | 1 | Complex | 1 | 0 | 0 | 3 |
| SCHI0125-P | 50-55 | F | 0 | 1 | 1 | 16 | 547 | 0.67 | 296 | 68 | 173 | 38 | - | 5.4 | 1 | Complex | 1 | 0 | 0 | 3 |
| SCHI0126-P | 70-75 | M | 0 | 1 | 1 | 15.7 | 191 | 0.11 | 183 | 60 | 581 | 34 | 7.3 | 9.2 | 1 | Complex | 1 | 0 | 0 | 2 |
| SCHI0127-P | 35-40 | F | 0 | 1 | 0 | 15.8 | 511 | 3.12 | 191 | 60 | 6920 | 44 | 7 | <0.3 | 1 | Simple | 0 | 1 | 0 | 3 |
| SCHI0128-P | 45-50 | M | 0 | 1 | 0 | 8.4 | 119 | 0.11 | 405 | 60 | 280 | 37 | 7.4 | 6.2 | 0 | Simple | 0 | 0 | 1 | 2 |
| SCHI0131-P | 80-85 | M | 0 | 1 | 1 | 10.2 | 39 | 7.72 | 201 | 61 | 2130 | 30 | - | <0.3 | 1 | Complex | 0 | 1 | 1 | 3 |
| SCHI0132-P | 20-25 | M | 0 | 1 | 0 | 12.9 | 229 | 1.42 | 161 | 68 | 1550 | 52 | 7 | 2.5 | 1 | Complex | 1 | 0 | 0 | 3 |
| SCHI0133-P | 25-29 | M | 0 | 1 | 1 | 27.6 | 97 | 0.28 | 275 | 70 | 897 | 48 | 6.8 | 2.4 | 1 | Complex | 1 | 0 | 0 | 3 |
| SCHI0134-P | 60-65 | F | 0 | 1 | 0 | 14.4 | 217 | 6.29 | 108 | 64 | 856 | 42 | 6.8 | <0.3 | 0 | Complex | 1 | 0 | 1 | 2 |
| SCHI0135-P | 70-75 | M | 0 | 1 | 0 | 13 | 246 | 0.99 | 264 | 64 | 514 | 48 | 7 | 3.6 | 0 | Complex | 1 | 0 | 0 | 3 |
| SCHI0136-P | 90-95 | M | 0 | 1 | 0 | 33.6 | 519 | 0.47 | 224 | 76 | 1680 | 53 | 6.6 | <0.3 | 1 | Complex | 1 | 0 | 0 | 3 |
| SCHI0140-P | 30-35 | M | 0 | 1 | 1 | 24.8 | 409 | 9.57 | 295 | 52 | 6210 | 49 | 6.9 | <0.3 | 1 | Complex | 1 | 1 | 0 | 3 |
| SCHI0141-P | 90-95 | M | 0 | 1 | 0 | 14.6 | 130 | 0.09 | 248 | 59 | 604 | 46 | 7.2 | 4.9 | 1 | Complex | 1 | 0 | 0 | 3 |
| SCHI0143-P | 65-70 | F | 1 | 1 | 0 | 12.1 | 119 | 2.13 | 358 | 49 | 874 | 23 | - | 3.8 | 1 | Complex | 1 | 0 | 0 | 2 |
| SCHI0148-P | 40-45 | F | 0 | 0 | 0 | 8.4 | 39 | 0.07 | 169 | 65 | 403 | 48 | 7.3 | 3.3 | 1 | Complex | 0 | 0 | 0 | 2 |
| SCHI0149-P | 70-75 | M | 0 | 0 | 0 | 8.5 | 81 | 0.05 | 286 | 67 | 1200 | 46 | 7.3 | 3.3 | 0 | Complex | 0 | 0 | 0 | 2 |
| SCHI0150-P | 75-80 | F | 1 | 1 | 0 | 20.1 | 270 | 4.33 | 298 | 56 | 1110 | 41 | 7.3 | 3.5 | 1 | Complex | 1 | 0 | 0 | 2 |
| SCHI0157-P | 30-35 | M | 0 | 1 | 0 | 12.3 | 99 | 0.08 | 169 | 69 | 727 | 58 | 6.6 | <0.3 | 1 | Complex | 1 | 0 | 0 | 3 |
| SCHI0158-P | 60-65 | M | 0 | 1 | 0 | 28.5 | 196 | 1.47 | 273 | 52 | 539 | 29 | 7.2 | 5.9 | 0 | Complex | 0 | 0 | 0 | 3 |
| SCHI0159-P | 70-75 | F | 0 | 1 | 0 | 11.6 | 135 | 0.82 | 477 | 54 | 292 | 29 | 7.5 | 6 | 1 | Complex | 0 | 0 | 0 | 2 |
| SCHI0160-P | 65-70 | M | 1 | 1 | 0 | 9.5 | 164 | 0.54 | 209 | 50 | 313 | 32 | 7.4 | 11.9 | 1 | Complex | 1 | 0 | 0 | 2 |
| SCHI0162-P | 55-60 | M | 0 | 1 | 0 | 9 | 106 | 0.16 | 291 | 65 | 610 | 48 | - | 3.9 | 1 | Complex | 1 | 1 | 0 | 2 |
| SCHI0166-P | 75-80 | M | 0 | 1 | 0 | 24.6 | 414 | 0.88 | 220 | 56 | 4990 | 49 | 6.6 | <0.3 | 0 | Complex | 1 | 0 | 0 | 3 |
| SCHI0163-P | 70-75 | M | 0 | 0 | 0 | 13.9 | 245 | 0.3 | 145 | 57 | 3990 | 46 | 7.0 | 3.5 | 1 | Complex | 1 | 0 | 0 | 3 |
| SCHI0169-P | 85-90 | M | 0 | 1 | 0 | 15.2 | 227 | 0.17 | 171 | 71 | 490 | 53 | - | 6.1 | 1 | Complex | 0 | 0 | 0 | 3 |
| SCHI0172-P | 20-25 | M | 0 | 1 | 1 | 16.4 | 171 | 0.17 | 110 | 49 | 17400 | 28 | - | <0.3 | 1 | Complex | 0 | 0 | 0 | 3 |
| SCHI0151-C | 70-75 | F | 0 | 0 | 0 | 10.5 | 83 | 0.14 | 236 | 62 | 393 | 45 | 7.3 | 10 | 0 | MPE | 0 | 0 | NA | 1 |
| SCHI0152-C | 80-85 | M | 0 | 0 | 0 | 10.3 | 90 | 0.19 | 160 | 62 | 81 | 39 | 7.3 | 4.9 | 0 | MPE | 0 | 0 | NA | 1 |
| SCHI0161-C | 85-90 | M | 0 | 0 | 0 | 10.5 | 26 | 0.11 | 172 | 72 | 232 | 41 | 7.3 | 7.9 | 0 | MPE | 0 | 0 | NA | 1 |
| SCHI0164-C | 75-80 | M | 0 | 0 | 0 | 5.2 | 124 | - | 250 | 62 | 427 | 37 | 7.3 | 4.6 | 0 | MPE | 0 | 0 | NA | 1 |
| SCHI0165-C | 80-85 | F | 0 | 0 | 0 | 15.2 | 29 | 0.26 | 194 | 72 | 99 | 39 | 7.4 | 6 | 0 | CCF | 0 | 0 | NA | 1 |
| SCHI0168-C | 40-45 | M | 0 | 0 | 0 | 9.6 | 7.3 | - | 287 | 45 | 287 | 45 | 7.4 | 5.2 | 0 | MPE | 0 | 0 | NA | 1 |
| SCHI0170-C | 60-65 | M | 0 | 0 | 0 | 4.5 | - | - | 182 | - | 91 | 46 | 7.5 | 7.8 | 0 | MPE | 0 | 0 | NA | 1 |
| SCHI0173-C | 70-75 | M | 0 | 0 | 0 | 6.9 | 84 | - | 391 | 61 | 672 | 39 | 7.4 | 6.3 | 0 | MPE | 0 | 0 | NA | 1 |
| SCHI0174-C | 75-80 | M | 1 | 0 | 0 | 6.7 | - | - | 222 | 67 | 245 | 44 | - | 9.2 | 0 | MPE | 0 | 0 | NA | 1 |
| SCHI0175-C | 85-90 | M | 0 | 0 | 0 | 4.3 | 5 | - | 193 | 70 | 88 | 45 | - | 6.2 | 0 | MPE | 0 | 0 | NA | 1 |

*Abbreviations:* CCF, Congestive Cardiac Failure; CTS, Cardiothoracic Surgical decortication; IPE, Intrapleural Enzyme therapy; MPE, Malignant Pleural Effusion; SIRS, Systemic Inflammatory Response Syndrome; LDH, Lactate dehydrogenase. 0=no, 1=yes.

^†^The -P suffix denotes participants suspected of having a pleural infection, whereas the -C suffix denotes participants suspected to have a non-infectious basis for their pleural effusion (i.e. negative controls). * 1) *Unlikely pleural infection:* Gram stain- or culture-negative with pH > 7.2, CRP < 100 mg/L, no fever, simple effusion, and a confirmed alternative diagnosis (i.e. heart failure, malignancy); 2) *Possible pleural infection:* Gram stain- or culture-negative with pH > 7.2 and 2 or more of the following: CRP > 100 mg/L; complex pleural fluid on imaging; fever (> 38.0 ^o^C); 3) *Definite or probable pleural infection:* Gram stain- and/or culture-positive pleural fluid; or pleural fluid pH < 7.2 and 2 or more of the following: serum C-reactive protein > 100 mg/L; complex pleural fluid on imaging; fever (> 38.0^o^C).
