## Supplementary material for "Evaluating the feasibility, sensitivity, and specificity of next-generation molecular methods for pleural infection diagnosis": Table S1

**Table S1. Antimicrobial therapy type, time to pleural fluid (PF) collection, and total duration in participants diagnosed with pleural infection, along with detected antimicrobial resistance (AMR) loci**

| **Participant ID** | **Antibiotics administered** | **Time prior to PF collection** | **AMR genes*** |
| --- | --- | --- | --- |
| *Participants diagnosed with pleural infection* | | | |
| SCHI0123-P | AMC, AZM, CRO, TZP | 64 h | Nil |
| SCHI0125-P | AZM, CRO, TZP | 22 h | Nil |
| SCHI0126-P | AMC | 96 h | Nil |
| SCHI0127-P | AZM, MXF | 15 h | *erm*(*A*) (streptogramin b resistance, CLIr, ERYr, LINr) |
| SCHI0128-P | CRO, TZP | 112 h | Nil |
| SCHI0131-P | AZM, TZP | 6 h | *cfxA3* (AMPr), *mef*(*A*) (ERYr, AZMr), *msr*(*D*) (AZMr, ERYr, telithromycin resistance, streptogramin b resistance), *tet*(*32*) and *tet*(*O*) (DOXr, MINr, TETr) |
| SCHI0132-P | AZM, CRO, TZP | 120 h | Nil |
| SCHI0133-P | CRO, TZP, VAN | 48 h | Nil |
| SCHI0134-P | TZP | 19 h | Nil |
| SCHI0135-P | TZP | 7 h | Nil |
| SCHI0136-P | TZP | 3 h | Nil |
| SCHI0140-P | AZM, TZP, VAN | 8 h | Nil |
| SCHI0141-P | AMC (oral for 5 days), TZP (IV for 72h) | 72 h | Nil |
| SCHI0143-P | CAZ | 21 days | Nil |
| SCHI0148-P | Nil | 0 h | Nil |
| SCHI0149-P | Nil | 0 h | Nil |
| SCHI0150-P | AZM, BPG, CRO | 102 h | Nil |
| SCHI0157-P | CAZ, DOX, metronidazole | 13 h | Nil |
| SCHI0158-P | AMC, FEP, TZP | 144 h | Nil |
| SCHI0159-P | AZM, BPG, CRO | 10 days | Nil |
| SCHI0160-P | TZP | 21 days | Nil |
| SCHI0162-P | AZM, BPG, CRO, TZP | 10 days | Nil |
| SCHI0163-P | AMC | 24 h | *aph(6)-Id* (STRr) |
| SCHI0166-P | AMC, BPG, DOX | 72 h | Nil |
| SCHI0169-P | AMX (then AMC), DOX | 94 h | Nil |
| SCHI0172-P | AMC, TZP | 36 h | *tet*(*M*) (DOXr, MINr, TETr) |

*Abbreviations:* AMC, amoxicillin-clavulanate; AMR, antimicrobial resistance; AMX, amoxicillin; AZM, azithromycin; AZMr, azithromycin resistance; BPG, benzathine penicillin G; CAZ, ceftazidime; CLIr, clindamycin resistance; CRO, ceftriaxone; DOX, doxycycline; DOXr, doxycycline resistance; ERYr, erythromycin resistance; FEP, cefepime; IV, intravenous; LINr, lincomycin resistance; MINr, minocycline resistance; MXF, moxifloxacin; STRr, streptomycin resistance; TETr, tetracycline resistance; TZP, piperacillin/tazobactam; VAN, vancomycin. *Identified from metagenomic data using ResFinder
